# Dietary sodium intake and kidney tubular protein abundance in urinary extracellular vesicles

**DOI:** 10.64898/2026.10.04.26364680

**Authors:** Sophie E. Claudel, Sanan Mahrokhian, Isabelle Hanna, Andrew J. Newman, Jenifer Brown, Ashish Verma, Sushrut S. Waikar, Richard J. Auchus, W. Hayes McDonald, Kevin L. Schey, Dungeng Peng, James Matthew Luther, Anand Vaidya

## Abstract

**Background:** Dietary sodium intake is a modifiable determinant of blood pressure, and impaired renal sodium handling is implicated in the pathogenesis of hypertension. Whether dietary sodium intake modulates renal tubular sodium transporter abundance among normotensive adults at risk for hypertension is unknown.

**Methods:** Forty-six normotensive adults with normal kidney function at risk of developing hypertension were prospectively enrolled to complete a very low sodium (LS) diet followed by a high sodium (HS) diet. Urinary extracellular vesicle (uEV) proteins were quantified from a 24-hour urine collection at the end of each diet. We compared normalized uEV protein concentrations between diets and tested whether diet modified uEV protein associations with serum aldosterone, urinary aldosterone, and serum cortisol-cortisone ratio under either diet using Wilcoxon rank sums and linear mixed effects models, respectively.

**Results:** Participants achieved median [IQR] urine sodium 13.2 [7.7-19.9] mEq/24h on the LS diet and 244.0 [197.1-297.0] mEq/24h on the HS diet. LS intake induced greater renin activity, greater plasma and urinary aldosterone, and lower urinary cortisol, when compared to HS intake. In parallel, NCC, ENaC, SGLT2, OXSR1, and Pendrin uEV concentrations were significantly higher during LS intake than HS intake (*p*<0.05 for all), whereas TRPV5 and AQP2 uEV concentrations were lower during LS intake than HS intake (*p*<0.001 for both).

**Conclusions:** Controlled dietary sodium modulation induced nephron-segment specific changes in uEV protein abundance. These findings demonstrate the ability of uEVs to serve as a non-invasive tool to probe *in vivo* renal sodium, water, and electrolyte handling.

## Introduction

Excessive dietary sodium intake is a principal modifiable determinant of blood pressure, and impaired renal sodium handling is implicated in the pathogenesis of salt-sensitive hypertension.^1^ Salt sensitivity of blood pressure is disproportionately common among individuals with obesity and insulin resistance, and may precede clinically evident hypertension by years.^2–6^ Yet measuring salt sensitivity in the clinic is challenging,^6^ and no clinically available test identifies tubular mechanisms of sodium handling in individuals with or at risk for hypertension. Clinicians therefore rely on antihypertensive therapies defined by the tubular targets they inhibit, while the sodium handling itself is only assessed in aggregate by surrogate markers.

Quantifying segment-specific tubular protein abundance in humans historically required invasive approaches such as kidney biopsy, or relied on indirect functional measures.^7,8^ Urinary extracellular vesicles (uEVs), which carry membrane-associated protein cargo shed from renal tubular epithelial cells, offer a non-invasive alternative.^7,9–12^ Aldosterone-responsive proteins, including ENaC subunits, are detectable in uEVs and can be used to study aldosterone-mediated sodium regulation.^8,13–15^ However, nearly all clinical uEV studies have been performed in patients with primary aldosteronism or established hypertension, where the physiology of sodium transport is already deranged.^16–20^ Whether uEV cargo is systematically altered by dietary sodium intake in normotensive individuals, and whether such changes relate to physiological variation in aldosterone, has not been examined. Establishing this relationship would determine whether uEVs can offer physiologically concordant, segment-specific insight into tubular sodium handling before hypertension develops and when it remains most modifiable.

We therefore studied variations in uEVs relevant to renal sodium and water handling among normotensive adults with cardiometabolic risk factors for hypertension during controlled and protocolized low– and high-sodium diets. We hypothesized that dietary sodium modulation would produce nephron-segment-specific changes in uEV protein abundance using a novel targeted mass spectrometry assay based on known renal physiology related to volume expansion, and that these changes would correlate with concurrent mineralocorticoid receptor activity. Two prior analyses of this cohort have demonstrated associations between circulating EVs and microvascular endothelial inflammatory markers,^21^ as well as urinary micro-RNA and renal inflammatory pathways.^22^ However, whether uEVs reflect *in vivo* changes in renal physiology during dietary sodium modulation among normotensive people has not been reported.

## Methods

### Study Population

The study population included participants prospectively recruited in Boston, MA between 2018 and 2023 (NCT03484130) for a longitudinal study of dynamic phenotyping to characterize subclinical aldosteronism.^23^ Participants were included according to the following criteria: age 35 to 70 years, an estimated glomerular filtration rate >60 mL/min per 1.73 m^2^, an untreated BP 120 to 135/75 to 85 mmHg, and at least 1 condition among body mass index ≥25 kg/m^2^, family history of hypertension before the age of 60 years in a parent or sibling, or diabetes with a hemoglobin A1c <9%. If systolic BP was 115 to 120 mm Hg or diastolic BP was 70 to 75 mmHg, the participants had to fulfill at least 2 criteria among those described above. Participants were excluded if they met any of the following conditions: hypertension, active malignancy, pregnancy, cardiovascular disease, glomerular filtration rate <60 mL/min per 1.73m^2^, untreated hyper– or hypothyroidism, or took anti-hypertensive medications, non-steroidal anti-inflammatory medications, or glucocorticoids. Participants completed a non-randomized crossover dietary intervention that was prepared and monitored by professional research dieticians.^23,24^ Professional dieticians prepared the low sodium (LS) diet first, which included prepared meals for 5 to 7 days designed to include ≈10 to 40 mEq/d of sodium (0.23–0.92 g/d) and ≈50– 75 mEq/d of potassium. Once the LS diet was completed, participants completed 5 to 7 days of the high sodium (HS) diet with a target >200 mEq/d (or 4.6 g/d) of sodium, maintaining ≈50 to 75 mEq/d of potassium. The high-sodium phase involved dietary counseling by professional dieticians to increase dietary sodium intake with sodium-rich foods and in addition sodium-rich broth packets when participants’ daily sodium intake was not considered high enough. A 24-hour urine home collection was obtained at the end of each dietary condition. All blood samples were collected in the morning at 8 am after an overnight fast. The study was approved by the Mass General Brigham Institutional Review Board; written informed consent was obtained from all participants.

For the purposes of this analysis, we excluded participants without uEV data or 24-hour urine sodium data due to incomplete urine collection. We additionally excluded participants with 24-hour urine sodium >50 mEq/24h following the LS diet or <150 mEq/24h following the HS diet, as these values suggest inadequate compliance with the dietary modification. A total of 59 participants had complete data for at least one diet condition (56 in the LS diet and 49 in the HS diet), with 46 participants meeting criteria under both diet conditions.

### Blood Pressure Measurement

Full study procedures have been previously described.^23–25^ Briefly, standardized, unattended in-office BPs were obtained after 30 minutes of seated rest and were measured in triplicate and averaged (Omron HEM-907XL; Omron Healthcare, Lake Forest, IL), following SPRINT trial protocol.^26^

### Laboratory Assays

Following each diet condition, morning (8-9am) seated and supine blood samples were assayed for plasma renin activity, plasma aldosterone, serum cortisone, serum cortisol, and serum electrolytes. Plasma renin activity was measured by angiotensin-I ELISA (IBL-International; Hamburg, Germany; inter-assay coefficient of variation (CV) = 7.6%–19.1%, intra-assay CV = 4.5%–11.4%; RRID:AB 3532145) after incubation at 37°C for 1.5 hours. Plasma aldosterone was measured by ELISA (IBL-America; inter-assay CV = 8.6%–9.4%, intra-assay CV = 3.9%– 9.7%, analytical sensitivity <5.7 pg/mL, assay dynamic range 5.7–1000 pg/mL; RRID:AB_2813725). Serum cortisol and cortisone were measured by LC-MS/MS.^27^ 24-hour urine collections were used to measure aldosterone by ELISA (IBL-America) to reflect the 24-hour urinary aldosterone.

### Urinary extracellular vesicle analysis

We isolated uEVs as previously described.^28,29^ Prior to uEV isolation, urine samples were spun at 5,000 g for 10 minutes and the supernatant was collected for uEV isolation. After centrifugation at 17,000 g, the supernatant was spun at 200,000 g for 1 hour and the uEV pellet was collected and suspended in 1X PBS buffer (pH 7.4). After trypsin digestion, target proteins were quantified by targeted multiple reaction monitoring (MRM) mass spectrometry on a ThermoScientific Vantage triple quadrupole LC-MS/MS instrument using isotopically labelled internal standards. To allow for efficient quantitation of this number of peptides, a chromatography time-scheduled MRM was performed on two injections using 1 µg per sample injection. Peptide target sequences are provided in **Table S1**. Peptides with standard curve R2 value <0.95 were excluded from analysis. The concentration of the uEV proteins were measured using sotopically labelled peptide standards for accurate quantification.^28,29^ To account for variability in vesicle excretion, isoloation yield, and urine dilution, uEV protein abundance was normalized to the exosomal tetraspanin CD9 to adjust for uEV protein content, Tamm-Horsfall depletion, and trypsin digest efficiency.^8,10,30^ For uEV proteins that had more than one subunit or peptide, we additionally created a combined protein variable by adding the concentrations of the individual subunits or peptides together.

### Statistical Analysis

We summarized continuous variables using medians and interquartile ranges (IQR) or means and standard deviations (SD), as appropriate. We log-transformed uEV protein concentration ratios, serum aldosterone concentration, urine aldosterone concentration, and serum cortisol-cortisone ratio prior to regression modeling to improve normality and stabilize variance.

To compare paired uEV protein:CD9 concentration ratios (hereafter referred to as uEV concentrations) between LS and HS diets, we performed Wilcoxon signed-rank tests within participants with measurements under both dietary conditions (N=46 matched pairs). As a complementary approach, we fit linear mixed-effects models using diet as a fixed effect and participant ID as a random intercept (*lme4* package). These models estimated overall diet-associated differences in uEV concentration while accounting for within-participant correlation, and allow for use of all available data (N=59 unique participants). Percent difference between the HS and LS diets was derived from the linear mixed effects model β coefficients.

We then categorized raw plasma aldosterone concentration, urine aldosterone excretion, and cortisol-cortisone ratio (a marker of 11-beta-hydroxysteroid dehydrogenase-2 (11β-HSD2) activity^31^) into tertiles; these were defined separately within each dietary sodium condition, such that tertile categories reflect relative aldosterone exposure within a given diet rather than absolute aldosterone concentrations across diets. Within each diet group, we summarized uEV concentrations across tertiles using median (IQR) and assessed differences across tertiles using the Kruskal–Wallis test. To evaluate monotonic trends across tertiles, we performed an ordinal trend test by modeling tertile as an ordered numeric variable in a linear regression framework. We visualized the distribution of log-transformed uEV concentrations across tertiles and diet conditions using boxplots with individual observations plotted.

Among participants with suppressed plasma renin activity (<1.0 ng/mL/hr) and elevated aldosterone (≥10 ng/dL) during the high-sodium condition, reflecting renin-independent aldosterone excess, we evaluated the correlation between uEV concentration and serum aldosterone concentration (Spearman’s rho).

To assess the association between uEV and serum aldosterone concentrations and assess for effect modification by dietary sodium intake, we fit linear mixed-effects models (*lme4* package), including an interaction term for diet and using all available data (N=59 participants). Diet-specific slopes for aldosterone were estimated using marginal trends (*emmeans* package), which accounts for covariance between fixed effects. All models included a random intercept for participant ID and were fit using restricted maximum likelihood for parameter estimation. Interaction p-values < 0.05 were considered evidence of diet-specific aldosterone regulation, acknowledging that this represents an exploratory threshold given multiple comparisons. Models were fit separately for each uEV protein, reflecting our *a priori* interest in uEV protein-specific relationships. Both uEV and the serum aldosterone concentrations were log-transformed, so coefficients represent the percent difference in uEV concentration per 1% difference in serum aldosterone concentration. Log-transformed exposures were mean-centered prior to modeling, such that the diet main effect represents the difference between dietary conditions at the mean serum aldosterone. The same modeling approach was then applied to urine aldosterone concentration and serum cortisol-cortisone ratio.

We then assessed participants’ salt sensitivity of blood pressure and assessed the correlation between salt sensitive blood pressure (SSBP) and changes in uEV concentration across HS and LS diets. SSBP was defined by subtracting the LS diet seated blood pressure from the HS diet seated blood pressure for each participant with data for both diet conditions. Salt sensitivity is indicated by blood pressure rising under the HS diet.

Analyses were performed in R software (version 4.5.2) and the *ggplot2* package was used to generate graphics. A large language model (Claude Opus 5) was used to debug and verify code. Analyses and study protocols were not blinded, and a formal power calculation was not conducted.

## Results

Participants followed the LS diet for a mean of 4.92 (SD 0.4) days and the HS diet for 5.4 (SD 0.8) days (**Table 1**). Blood pressure, heart rate, and serum potassium were similar under both dietary conditions. Multiple physiologically expected changes were observed between the LS and HS dietary conditions, including, lower serum chloride under LS (100.5 (SD 1.8) mg/dl vs. 103.6 (SD 2.0) mg/dl), lower mean eGFR with LS intake (86.4 (SD 12.0) ml/min per 1.73m^2^ vs. 94.4 (SD 11.6) ml/min per 1.73m^2^), higher plasma renin activity and aldosterone were with LS intake, while serum cortisol-cortisone ratio was similar (**Figure 1**). With LS intake, median [interquartile range (IQR)] 24-hour urine studies showed higher aldosterone excretion (17.1 [IQR 11.0-28.0] μg/24h vs. 4.5 [IQR 3.3-5.5] μg/24h), lower cortisol excretion (12.9 [IQR 8.3-17.9] μg/24h vs. 25.1 [IQR 15.4-34.3] μg/24h), lower sodium (13.2 [IQR 7.7-19.9] mEq/24h vs. 244.0 [IQR 197.1-297.0] mEq/24h) and lower potassium (40.7 [IQR 31.5-54.2] mEq/24h vs. 86.9 [IQR 61.2-120.8] mEq/24h).

**Figure 1.**
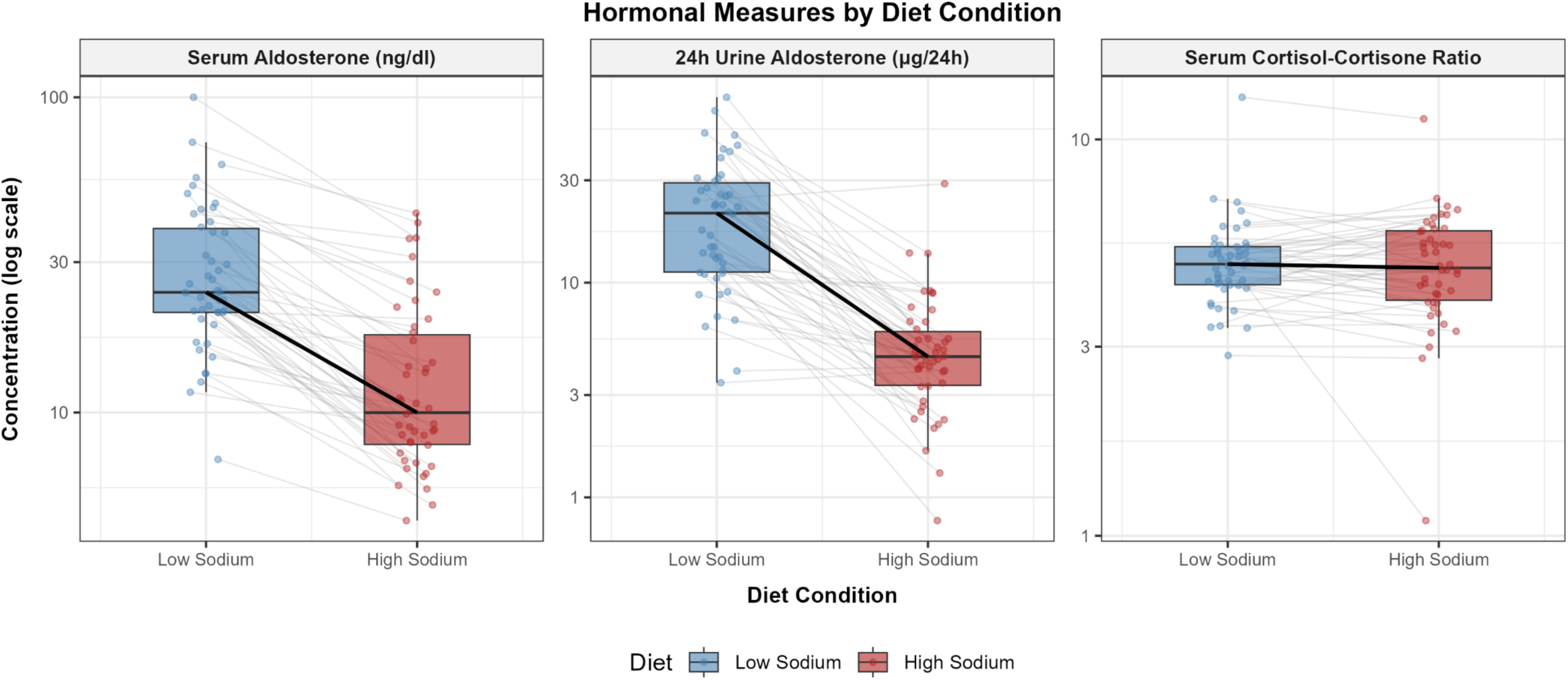
Boxplots summarizing the distribution of serum aldosterone, urine aldosterone, and serum cortisol-cortisone ratio across diet conditions, among matched pairs (N=46). Boxplots show the median (center line), interquartile range (box), and whiskers extending to the most extreme values within 1.5x the interquartile range. Individual observations are overlaid. Thin gray lines connect paired values from the same participant. Solid black line connects the median across all participants.

**Table 1.**
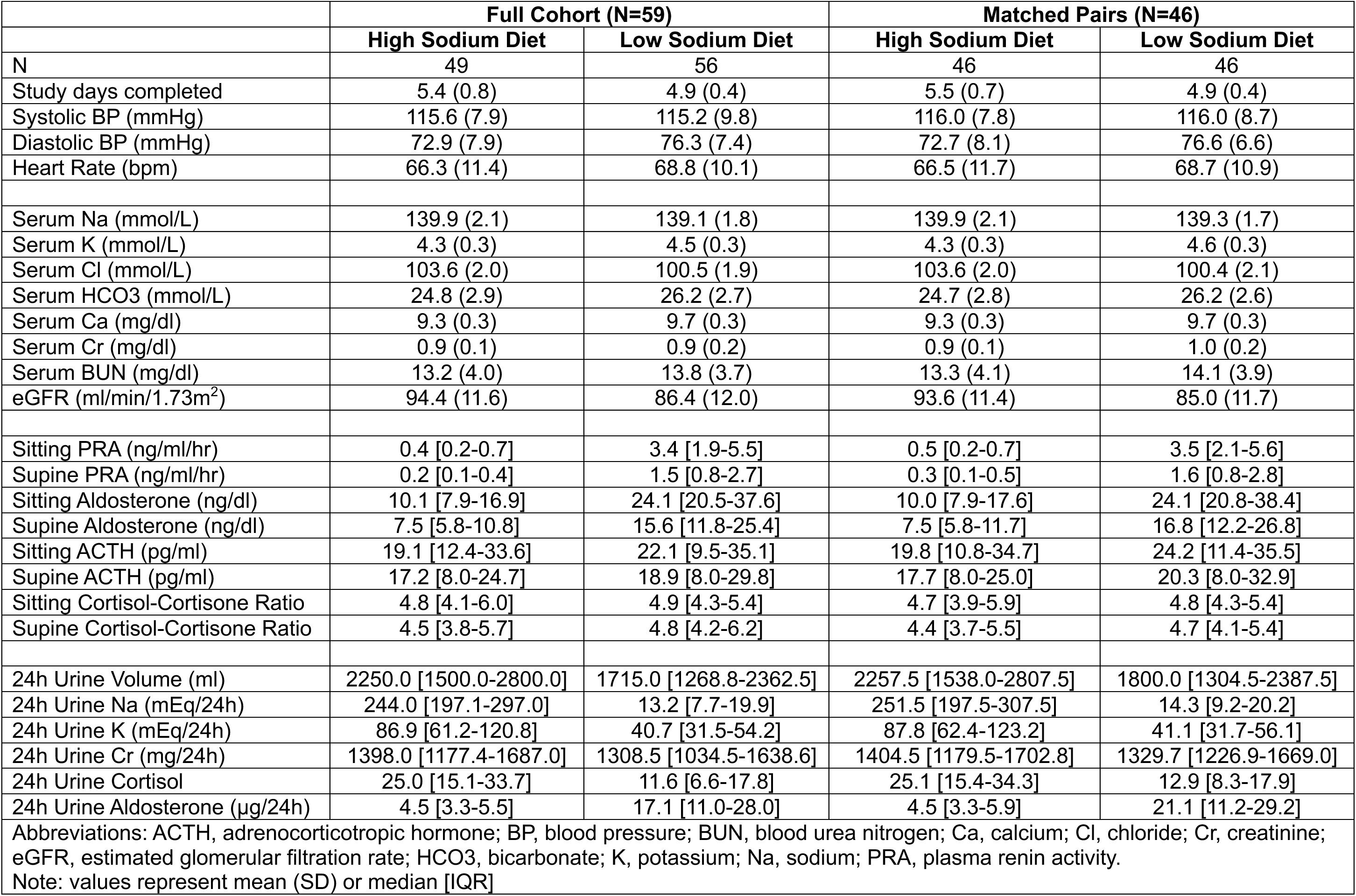
Population Characteristics by Dietary Sodium Intake.

| **Table 1. Population Characteristics by Dietary Sodium Intake** |  |  |  |  |
| --- | --- | --- | --- | --- |
|  | **Full Cohort (N=59)** |  | **Matched Pairs (N=46)** |  |
|  | **High Sodium Diet** | **Low Sodium Diet** | **High Sodium Diet** | **Low Sodium Diet** |
| N | 49 | 56 | 46 | 46 |
| Study days completed | 5.4 (0.8) | 4.9 (0.4) | 5.5 (0.7) | 4.9 (0.4) |
| Systolic BP (mmHg) | 115.6 (7.9) | 115.2 (9.8) | 116.0 (7.8) | 116.0 (8.7) |
| Diastolic BP (mmHg) | 72.9 (7.9) | 76.3 (7.4) | 72.7 (8.1) | 76.6 (6.6) |
| Heart Rate (bpm) | 66.3 (11.4) | 68.8 (10.1) | 66.5 (11.7) | 68.7 (10.9) |
| Serum Na (mmol/L) | 139.9 (2.1) | 139.1 (1.8) | 139.9 (2.1) | 139.3 (1.7) |
| Serum K (mmol/L) | 4.3 (0.3) | 4.5 (0.3) | 4.3 (0.3) | 4.6 (0.3) |
| Serum Cl (mmol/L) | 103.6 (2.0) | 100.5 (1.9) | 103.6 (2.0) | 100.4 (2.1) |
| Serum HCO<sub>3</sub> (mmol/L) | 24.8 (2.9) | 26.2 (2.7) | 24.7 (2.8) | 26.2 (2.6) |
| Serum Ca (mg/dl) | 9.3 (0.3) | 9.7 (0.3) | 9.3 (0.3) | 9.7 (0.3) |
| Serum Cr (mg/dl) | 0.9 (0.1) | 0.9 (0.2) | 0.9 (0.1) | 1.0 (0.2) |
| Serum BUN (mg/dl) | 13.2 (4.0) | 13.8 (3.7) | 13.3 (4.1) | 14.1 (3.9) |
| eGFR (ml/min/1.73m<sup>2</sup>) | 94.4 (11.6) | 86.4 (12.0) | 93.6 (11.4) | 85.0 (11.7) |
| Sitting PRA (ng/ml/hr) | 0.4 [0.2-0.7] | 3.4 [1.9-5.5] | 0.5 [0.2-0.7] | 3.5 [2.1-5.6] |
| Supine PRA (ng/ml/hr) | 0.2 [0.1-0.4] | 1.5 [0.8-2.7] | 0.3 [0.1-0.5] | 1.6 [0.8-2.8] |
| Sitting Aldosterone (ng/dl) | 10.1 [7.9-16.9] | 24.1 [20.5-37.6] | 10.0 [7.9-17.6] | 24.1 [20.8-38.4] |
| Supine Aldosterone (ng/dl) | 7.5 [5.8-10.8] | 15.6 [11.8-25.4] | 7.5 [5.8-11.7] | 16.8 [12.2-26.8] |
| Sitting ACTH (pg/ml) | 19.1 [12.4-33.6] | 22.1 [9.5-35.1] | 19.8 [10.8-34.7] | 24.2 [11.4-35.5] |
| Supine ACTH (pg/ml) | 17.2 [8.0-24.7] | 18.9 [8.0-29.8] | 17.7 [8.0-25.0] | 20.3 [8.0-32.9] |
| Sitting Cortisol-Cortisone Ratio | 4.8 [4.1-6.0] | 4.9 [4.3-5.4] | 4.7 [3.9-5.9] | 4.8 [4.3-5.4] |
| Supine Cortisol-Cortisone Ratio | 4.5 [3.8-5.7] | 4.8 [4.2-6.2] | 4.4 [3.7-5.5] | 4.7 [4.1-5.4] |
| 24h Urine Volume (ml) | 2250.0 [1500.0-2800.0] | 1715.0 [1268.8-2362.5] | 2257.5 [1538.0-2807.5] | 1800.0 [1304.5-2387.5] |
| 24h Urine Na (mEq/24h) | 244.0 [197.1-297.0] | 13.2 [7.7-19.9] | 251.5 [197.5-307.5] | 14.3 [9.2-20.2] |
| 24h Urine K (mEq/24h) | 86.9 [61.2-120.8] | 40.7 [31.5-54.2] | 87.8 [62.4-123.2] | 41.1 [31.7-56.1] |
| 24h Urine Cr (mg/24h) | 1398.0 [1177.4-1687.0] | 1308.5 [1034.5-1638.6] | 1404.5 [1179.5-1702.8] | 1329.7 [1226.9-1669.0] |
| 24h Urine Cortisol | 25.0 [15.1-33.7] | 11.6 [6.6-17.8] | 25.1 [15.4-34.3] | 12.9 [8.3-17.9] |
| 24h Urine Aldosterone (µg/24h) | 4.5 [3.3-5.5] | 17.1 [11.0-28.0] | 4.5 [3.3-5.9] | 21.1 [11.2-29.2] |
| Abbreviations: ACTH, adrenocorticotrophic hormone; BP, blood pressure; BUN, blood urea nitrogen; Ca, calcium; Cl, chloride; Cr, creatinine; eGFR, estimated glomerular filtration rate; HCO<sub>3</sub>, bicarbonate; K, potassium; Na, sodium; PRA, plasma renin activity. |  |  |  |  |
| Note: values represent mean (SD) or median [IQR] |  |  |  |  |

uEV concentrations differed between HS and LS intake (**Table 2**, **Figure 2, Figure S1**). NCC, ENaC subunits, SGLT2, OXSR1, and Pendrin were all lower during HS intake than LS intake (Wilcoxon *p*<0.01 for all). The percent differences based on the linear mixed models showed lower concentrations in HS compared to LS: NCC percent difference –45.3% (95% CI – 58.6 to –27.7, *p*=0.0001), ENaC subunits combined –35.2% (95% CI –55.5 to –5.5, *p*=0.03), OXSR1 –30.1% (95% CI –48.5 to –5.2, *p*=0.03), and Pendrin –73.0% (95% CI –81.2 to –61.1, *p*<0.0001) lower during HS than LS.

**Figure 2.**
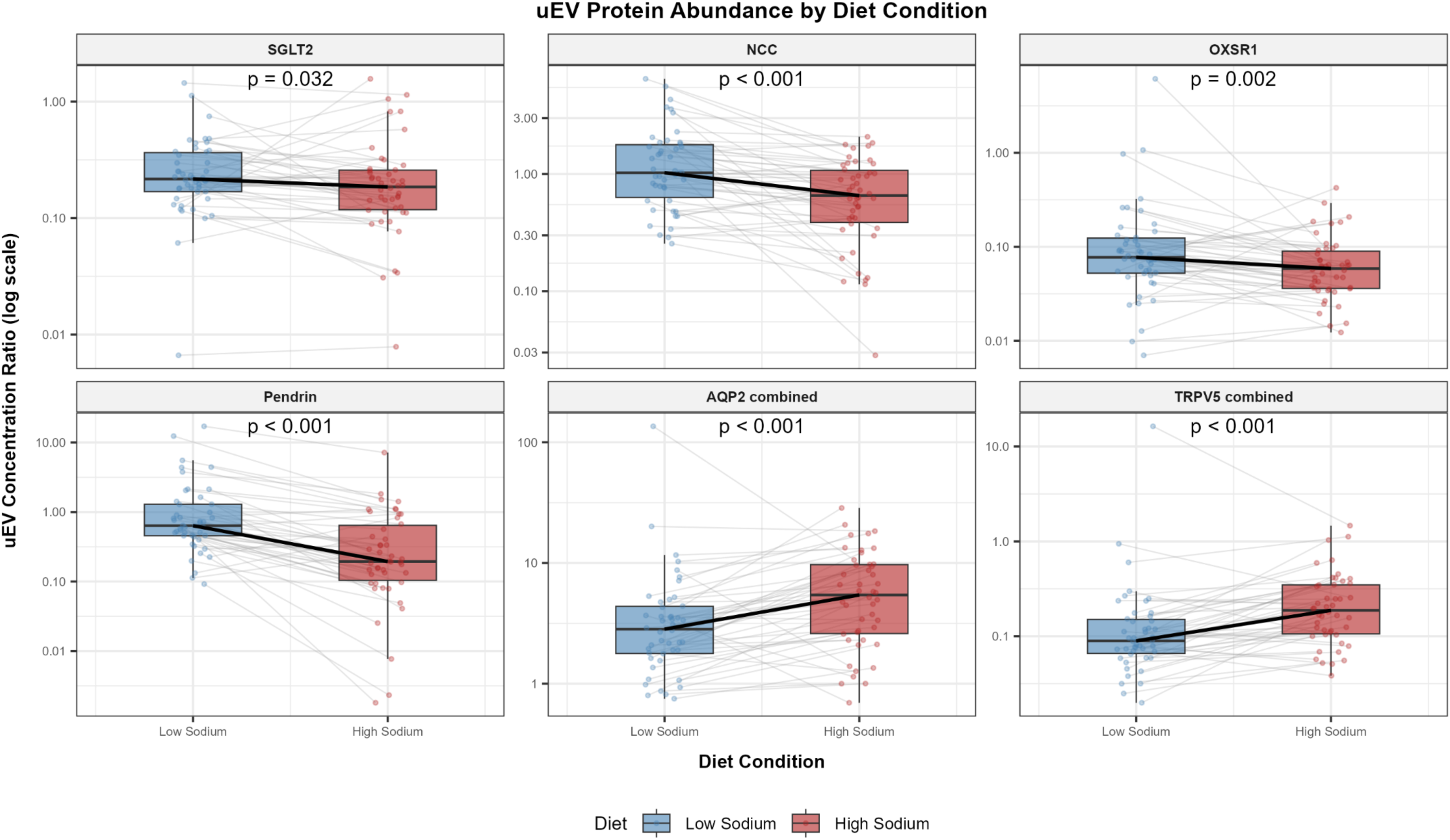
Boxplots summarizing the distribution of uEV concentration by diet for SGLT2, NCC, Pendrin, AQP2, and TRPV5, among matched pairs (N=46). Boxplots show the median (center line), interquartile range (box), and whiskers extending to the most extreme values within 1.5x the interquartile range. Individual observations are overlaid. Data are displayed on a log_10_ scale. Thin gray lines connect paired values from the same participant. Solid black line connects the median across all participants. P-values from Wilcoxon signed rank.

**Table 2.** Urine EV Concentration by Dietary Sodium Intake.

|  | **High Sodium Diet**<br>Median [IQR] | **Low Sodium Diet**<br>Median [IQR] | **Median Absolute**<br>**Difference (HS-LS) \*** | **p-value<sup>†</sup>** | **% Difference<sup>‡</sup>** | **p-value<sup>§</sup>** |
| --- | --- | --- | --- | --- | --- | --- |
| NCC | 0.73 [0.42-1.22] | 1.05 [0.73-1.82] | -0.38 (-0.81, -0.22) | <0.0001 | -45.3 (-58.6, -27.7) | 0.0001 |
| ENaC-α | 0.02 [0.01-0.02] | 0.02 [0.01-0.03] | -0.001 (-0.004, 0.004) | 0.71 | -20.5 (-38.4, 2.7) | 0.09 |
| ENaC-β | 0.01 [0.01-0.02] | 0.01 [0.01-0.02] | -0.003 (-0.005, 0.0003) | 0.07 | -30.6 (-52.9, 2.2) | 0.07 |
| ENaC-γ | 0.02 [0.01-0.03] | 0.02 [0.01-0.04] | -0.006 (-0.01, -0.002) | 0.01 | -35.5 (-60.2, 4.8) | 0.08 |
| ENaC combined | 0.05 [0.02-0.07] | 0.06 [0.03-0.09] | -0.01 (-0.02, -0.003) | 0.009 | -35.2 (-55.5, -5.5) | 0.03 |
| NKCC2 | 0.01 [0.00-0.01] | 0.01 [0.00-0.01] | 0.001 (-0.0008, 0.003) | 0.42 | 9.1 (-24.8, 58.5) | 0.65 |
| NKCC1 | 0.80 [0.46-1.27] | 0.81 [0.63-1.38] | -0.11 (-0.31, 0.08) | 0.36 | -21.4 (-41.1, 4.9) | 0.11 |
| NKCC combined | 0.81 [0.46-1.27] | 0.81 [0.64-1.39] | -0.11 (-0.29, 0.09) | 0.39 | -20.0 (-39.6, 5.9) | 0.13 |
| TRPV5_1 | 0.10 [0.06-0.20] | 0.04 [0.03-0.08] | 0.05 (0.02, 0.07) | <0.0001 | 95.5 (52.3, 150.9) | <0.0001 |
| TRPV5_2 | 0.09 [0.05-0.18] | 0.05 [0.03-0.08] | 0.04 (0.02, 0.07) | <0.0001 | 71.2 (26.1, 132.4) | 0.001 |
| TRPV5 combined | 0.20 [0.11-0.38] | 0.09 [0.07-0.17] | 0.09 (0.04, 0.15) | <0.0001 | 84.0 (40.5, 141.0) | 0.0001 |
| TBC1D4_1 | 0.01 [0.004-0.01] | 0.01 [0.005-0.02] | 0.0001 (-0.002, 0.003) | 0.87 | -8.0 (-34.0, 28.4) | 0.63 |
| TBC1D4_2 | 0.005 [0.003-0.02] | 0.01 [0.004-0.03] | -0.002 (-0.01, 0.0008) | 0.17 | -34.0 (-58.0, 3.5) | 0.08 |
| TBC1D4 combined | 0.01 [0.01-0.03] | 0.02 [0.01-0.05] | -0.002 (-0.01, 0.003) | 0.30 | -23.9 (-47.9, 11.2) | 0.17 |
| AQP2_1 | 2.04 [1.15-3.83] | 0.93 [0.61-1.69] | 0.93 (0.46, 1.49) | <0.0001 | 70.5 (26.6, 129.7) | 0.001 |
| AQP2_2 | 2.98 [1.25-5.46] | 1.25 [0.82-2.62] | 1.57 (0.82, 2.41) | <0.0001 | 78.0 (39.5, 127.0) | <0.0001 |
| AQP2 combined | 5.79 [2.67-10.62] | 2.24 [1.61-4.33] | 2.39 (1.36, 3.95) | <0.0001 | 79.9 (39.3, 132.3) | <0.0001 |
| SGLT2 | 0.19 [0.12-0.28] | 0.22 [0.17-0.39] | -0.05 (-0.09, -0.004) | 0.03 | -18.0 (-36.0, 5.1) | 0.12 |
| OXSR1 | 0.06 [0.04-0.10] | 0.08 [0.05-0.15] | -0.03 (-0.05, -0.01) | 0.002 | -30.1 (-48.5, -5.2) | 0.03 |
| KCNJ1 | 0.07 [0.04-0.15] | 0.08 [0.06-0.17] | -0.0004 (-0.02, 0.02) | 0.95 | -21.6 (-48.2, 18.7) | 0.26 |
| SGK1 | 0.01 [0.01-0.02] | 0.01 [0.01-0.02] | 0.0002 (-0.003, 0.006) | 0.97 | -7.3 (-32.1, 26.6) | 0.64 |
| Pendrin | 0.21 [0.13-0.83] | 0.65 [0.46-1.33] | -0.52 (-0.83, -0.37) | <0.0001 | -73.0 (-81.2, -61.1) | <0.0001 |
| Cullin3 | 0.03 [0.01-0.06] | 0.03 [0.02-0.06] | -0.002 (-0.01, 0.005) | 0.51 | -14.3 (-39.4, 21.2) | 0.39 |
| Abbreviations: CI, confidence interval; HS, high sodium diet; IQR, interquartile range; LS, low sodium diet.<br>Note: values are represented as median [IQR] within diet conditions, using all available data (LS N=56, HS N=49). uEV concentrations are normalized to CD9.<br>* Median (95% CI) of within-person differences for N=46 matched pairs.<br><sup>†</sup> Wilcoxon signed rank (N=46 matched pairs)<br><sup>‡</sup> Percent difference in geometric means, HS relative to LS, back-transformed from log-scale model coefficients as $(\exp(\beta)-1)*100$ from the linear mixed model. Confidence intervals are therefore asymmetric on the percentage scale.<br><sup>§</sup> Linear mixed model (LS N=56, HS N=49) |  |  |  |  |  |  |

In contrast, TRPV5, TBC1D4, and AQP2 were higher during HS intake than LS intake (Wilcoxon *p*<0.001 for all). The percent differences between HS and LS based on the linear mixed models were consistent with these findings for TRPV5 and AQP2: TRPV5 subunits combined percent difference –84.0% (95% CI –141.0 to –40.5, *p*=0.0001) and AQP2 subunits and isomers combined –79.9% (95% CI –132.3 to –39.3, *p*<0.0001).

Among participants with data in both diet conditions, systolic blood pressure rose by more than 10 mmHg with sodium loading in 8.7% (N=4) of participants, fell by more than 10 mmHg in 10.9% (N=5), and changed by 10 mmHg or less in 80.4% (N=37). The median change in systolic blood pressure was –1.0 mmHg [IQR –5.0 to 4.0]. There were no strong correlations between blood pressure changes and uEV concentration changes across the two diet conditions (**Figure 3, Figure S2**).

**Figure 3.**
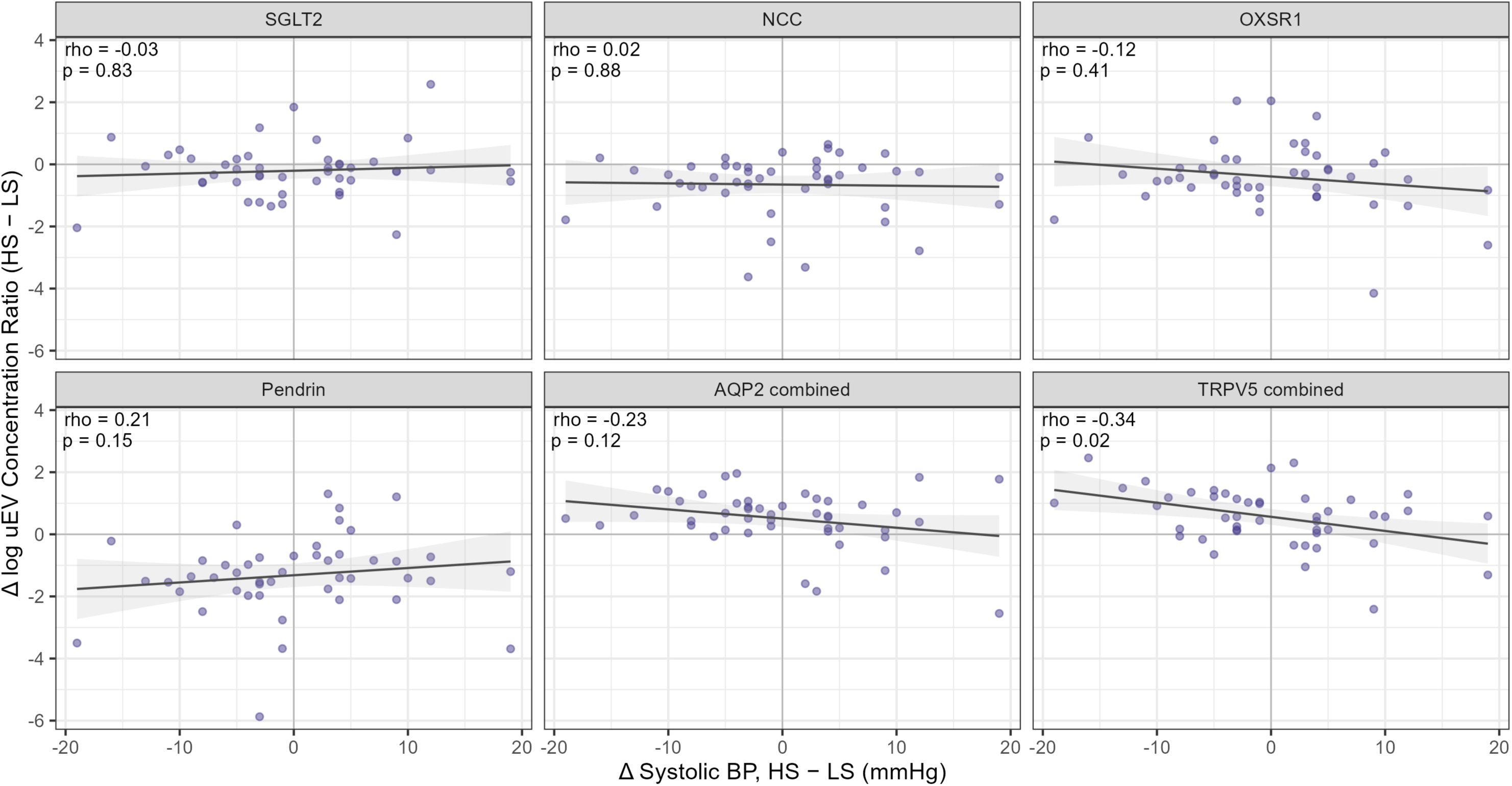
Scatterplots summarizing the correlation between change in uEV concentration and change in systolic blood pressure across diet conditions, among matched pairs (N=46). Both differences are calculated as high sodium minus low sodium, such that positive values indicate a higher value during high sodium intake. Change in uEV concentration is expressed as the difference in log-transformed values. Scatterplots show individual observations. The trend line is the ordinary least-squares regression fit and the shaded band is the 95% confidence interval around the fitted line. Spearman’s rho and the associated p-value are overlaid in each panel.

There were no clear monotonic relationships between plasma aldosterone, urine aldosterone, or serum cortisol-cortisone ratio and uEV concentrations under either diet condition (**Figures S3-5, Tables S2-4**). Few associations emerged when examining diet-specific slopes for the association between each uEV protein and the three hormonal markers (**Table 3**).

**Table 3.** Association between hormonal regulators of mineralocorticoid receptor activity and uEV concentration, with effect modification by dietary sodium intake (log-log models).

|  | Serum Aldosterone |  |  | Urine Aldosterone |  |  | Urine Cortisol-Cortisone Ratio |  |  |
| --- | --- | --- | --- | --- | --- | --- | --- | --- | --- |
|  | HS Diet<br>β (95% CI) | LS Diet<br>β (95% CI) | p-int | HS Diet<br>β (95% CI) | LS Diet<br>β (95% CI) | p-int | HS Diet<br>β (95% CI) | LS Diet<br>β (95% CI) | p-int |
| NCC | -0.03 (-0.49, 0.43) | -0.03 (-0.51, 0.46) | 0.99 | -0.27 (-0.64, 0.10) | -0.20 (-0.50, 0.11) | 0.73 | 0.31 (-0.45, 1.06) | 0.05 (-0.91, 1.01) | 0.63 |
| ENaC-α | 0.25 (-0.19, 0.68) | 0.51 (0.04, 0.98) | 0.27 | -0.01 (-0.37, 0.35) | -0.14 (-0.44, 0.16) | 0.53 | -0.33 (-1.05, 0.38) | -0.52 (-1.44, 0.41) | 0.71 |
| ENaC-β | -0.08 (-0.63, 0.47) | 0.19 (-0.38, 0.77) | 0.46 | -0.24 (-0.70, 0.21) | -0.02 (-0.38, 0.35) | 0.43 | 0.64 (-0.30, 1.57) | 0.06 (-1.11, 1.23) | 0.42 |
| ENaC-γ | -0.38 (-1.22, 0.46) | -0.10 (-0.99, 0.80) | 0.56 | -0.46 (-1.13, 0.21) | -0.12 (-0.68, 0.43) | 0.40 | -0.05 (-1.27, 1.18) | -0.48 (-2.03, 1.08) | 0.63 |
| ENaC combined | -0.15 (-0.83, 0.52) | 0.14 (-0.59, 0.87) | 0.43 | -0.36 (-0.90, 0.18) | -0.12 (-0.56, 0.33) | 0.44 | -0.02 (-0.97, 0.93) | -0.71 (-1.93, 0.51) | 0.32 |
| NKCC2 | -0.07 (-0.74, 0.61) | 0.02 (-0.70, 0.75) | 0.81 | -0.51 (-1.05, 0.02) | -0.36 (-0.80, 0.08) | 0.63 | 0.26 (-0.81, 1.32) | -0.85 (-2.25, 0.54) | 0.13 |
| NKCC1 | -0.43 (-0.96, 0.10) | -0.25 (-0.82, 0.32) | 0.53 | -0.54 (-0.95, -0.13) | -0.16 (-0.51, 0.18) | 0.12 | 0.78 (-0.07, 1.62) | -0.14 (-1.24, 0.96) | 0.12 |
| NKCC combined | -0.43 (-0.95, 0.09) | -0.27 (-0.83, 0.28) | 0.58 | -0.56 (-0.96, -0.15) | -0.16 (-0.50, 0.17) | 0.10 | 0.79 (-0.02, 1.61) | -0.14 (-1.21, 0.92) | 0.10 |
| TRPV5_1 | -0.17 (-0.64, 0.30) | -0.11 (-0.62, 0.40) | 0.82 | -0.14 (-0.51, 0.22) | -0.12 (-0.43, 0.19) | 0.90 | 0.28 (-0.48, 1.04) | 0.29 (-0.72, 1.29) | 0.99 |
| TRPV5_2 | -0.19 (-0.72, 0.34) | -0.18 (-0.75, 0.39) | 0.98 | -0.45 (-0.87, -0.03) | -0.15 (-0.49, 0.20) | 0.22 | 0.92 (0.06, 1.78) | 0.35 (-0.77, 1.47) | 0.34 |
| TRPV5 combined | -0.20 (-0.69, 0.29) | -0.17 (-0.70, 0.36) | 0.90 | -0.30 (-0.69, 0.08) | -0.14 (-0.47, 0.18) | 0.48 | 0.58 (-0.22, 1.37) | 0.30 (-0.74, 1.35) | 0.62 |
| TBC1D4_1 | -0.14 (-0.66, 0.38) | -0.11 (-0.65, 0.43) | 0.93 | -0.19 (-0.61, 0.23) | -0.20 (-0.54, 0.14) | 0.97 | 0.36 (-0.49, 1.20) | -0.11 (-1.19, 0.96) | 0.45 |
| TBC1D4_2 | -0.24 (-0.99, 0.51) | -0.27 (-1.06, 0.53) | 0.96 | -0.62 (-1.22, -0.02) | -0.14 (-0.63, 0.35) | 0.18 | 1.12 (-0.06, 2.30) | 0.62 (-0.89, 2.14) | 0.55 |
| TBC1D4 combined | -0.23 (-0.84, 0.38) | -0.19 (-0.83, 0.45) | 0.92 | -0.46 (-0.95, 0.03) | -0.12 (-0.51, 0.28) | 0.25 | 0.62 (-0.34, 1.58) | 0.33 (-0.89, 1.56) | 0.68 |
| AQP2_1 | -0.20 (-0.68, 0.28) | 0.04 (-0.46, 0.55) | 0.41 | -0.10 (-0.50, 0.29) | 0.08 (-0.24, 0.40) | 0.45 | 0.44 (-0.35, 1.23) | -0.57 (-1.58, 0.44) | 0.08 |
| AQP2_2 | 0.01 (-0.43, 0.45) | 0.19 (-0.28, 0.67) | 0.45 | 0.22 (-0.13, 0.57) | 0.22 (-0.08, 0.51) | 0.99 | 0.38 (-0.34, 1.11) | -0.17 (-1.12, 0.78) | 0.26 |
| AQP2 combined | -0.18 (-0.61, 0.25) | 0.01 (-0.45, 0.47) | 0.45 | 0.11 (-0.25, 0.46) | 0.13 (-0.16, 0.42) | 0.90 | 0.42 (-0.30, 1.13) | -0.23 (-1.15, 0.69) | 0.20 |
| SGLT2 | -0.49 (-0.92, -0.07) | -0.01 (-0.46, 0.45) | 0.04 | -0.44 (-0.79, -0.10) | -0.03 (-0.31, 0.25) | 0.04 | 0.45 (-0.24, 1.15) | 0.49 (-0.40, 1.39) | 0.93 |
| OXSR1 | -0.12 (-0.62, 0.38) | 0.28 (-0.25, 0.81) | 0.18 | -0.18 (-0.58, 0.23) | 0.09 (-0.25, 0.42) | 0.28 | 0.42 (-0.41, 1.25) | 0.08 (-0.97, 1.14) | 0.57 |
| KCNJ1 | -0.64 (-1.33, 0.06) | -0.38 (-1.12, 0.37) | 0.52 | -0.64 (-1.20, -0.08) | -0.36 (-0.82, 0.09) | 0.41 | -0.18 (-1.33, 0.97) | -0.08 (-1.56, 1.40) | 0.90 |
| SGK1 | 0.15 (-0.35, 0.65) | -0.37 (-0.90, 0.16) | 0.08 | -0.35 (-0.75, 0.06) | -0.38 (-0.71, -0.05) | 0.90 | 0.36 (-0.47, 1.18) | 0.45 (-0.60, 1.51) | 0.87 |
| Pendrin | 0.01 (-0.64, 0.65) | -0.11 (-0.80, 0.57) | 0.74 | -0.07 (-0.58, 0.44) | 0.07 (-0.36, 0.49) | 0.65 | -0.69 (-1.74, 0.35) | -0.48 (-1.84, 0.88) | 0.77 |
| Cullin3 | 0.09 (-0.56, 0.73) | -0.15 (-0.85, 0.54) | 0.49 | -0.26 (-0.77, 0.25) | -0.38 (-0.80, 0.05) | 0.69 | -0.09 (-1.13, 0.95) | -0.22 (-1.58, 1.14) | 0.85 |

Estimates derived from linear mixed-effects models with random intercept for participant and diet interaction. uEV concentration normalized to CD9. β represents a percent change in uEV per 1% change in aldosterone. Abbreviations: CI, confidence interval; HS, high sodium; LS, low sodium; int, interaction.

## Discussion

In this study of normotensive adults at risk for hypertension, we found that dietary sodium intake alters *in vivo* urinary tubular protein abundance across multiple nephron segments. The directionality of change was consistent with expected renal physiology, demonstrating that uEVs provide a real-time reflection of human physiology during dietary sodium interventions. Our findings complement existing literature on short-term physiologic interventions and changes in circulating EV concentrations.^15,21,22,32^ While few studies have used uEVs to examine normal tubular physiology or dynamic sodium handling, one study of 17 healthy Danish young men identified increased *SLC12A3* mRNA (which encodes NCC) when isolating uEVs under low sodium intake, compared to high sodium intake,^33^ though did not observe differences in absolute NCC or ENaC uEV abundance.^34^ Together with our findings, this growing body of literature highlights the emerging role of EVs in probing *in vivo* renal physiology.

The differences we observed in physiologic renin-angiotensin-aldosterone system activation indicate that the dietary modification protocol successfully maximizing aldosterone production under LS intake and suppressed aldosterone production under HS intake. Modulating aldosterone production using dietary sodium resulted in substantial shifts in the uEV profiles concordant with expected physiology. During HS intake, aldosterone suppression reduces sodium reabsorption via ENaC and NCC, increasing urinary sodium excretion.^35^ Concurrently, vasopressin-mediated AQP2 upregulation maintains water reabsorption, allowing the kidney to excrete high sodium loads in concentrated urine and minimize obligatory water loss.^36^ This prevents osmotic diuresis and volume depletion during sodium excretion, while also maintaining plasma osmolality.^37^ Given the similarity in blood pressure between the HS and LS intake conditions in our study, blood pressure alone is unlikely the driving force between the differential uEV concentrations between the two conditions. Additionally, the change in eGFR between the two diets likely reflects the changes in intravascular volume and glomerular perfusion pressure rather than a true change in kidney function. Our findings therefore support uEV profiling as a tool to interrogate real-time functional tubular responses to physiological stimuli.

Our findings complement preclinical research demonstrating diet-induced changes to renal sodium transporters. Under LS intake, the proximal tubule rapidly increases reabsorption of filtered sodium through the sodium-hydrogen exchanger (NHE3), sodium-phosphate (Na/Pi) cotransporter, and SGLT2 cotransporter, and the opposite holds for HS intake.^38–42^ We observed higher SGLT2 uEV concentration under LS intake. NCC is also strongly sodium and potassium responsive and is upregulated under LS and low potassium intake.^43–45^ NCC activity is also determined by distal chloride delivery and the activity of the with-no-lysine (WNK) kinase in the distal convoluted tubule, which is closely associated with the potassium-mediated regulatory control of NCC.^43,44^. We observed higher NCC uEV concentration during the LS intake, as anticipated, despite stable dietary potassium intake. As WNK kinases activate NCC via SPAK/OXSR1-mediated phosphorylation, our finding of higher OXSR1 uEV concentration during LS intake is directionally concordant with the observed change in NCC uEV concentration. Similarly, Pendrin, a chloride-bicarbonate exchanger on the apical membrane of the cortical collecting duct,^46,47^ was more abundant with LS intake in our study, which is consistent with a normal physiological response to volume depletion and low distal luminal chloride delivery.^47,48^ Although LS intake is also known to increase NKCC2 abundance to increase sodium chloride reabsorption and decrease distal sodium chloride delivery,^38,42^ we did not observe differences in NKCC2 uEV concentration between the diets. This may be due to methodological challenges in measuring very low-abundance proteins rather than a true physiologic response to LS intake. Similarly, although there was slight increase in the combined ENaC subunit concentration under LS intake, which is physiologically concordant,^42^ the methodological challenge of measuring the ENaC uEVs may have led to underestimation of the physiologic effect.

Given that direct measurement of tubular transporter abundance in humans is not currently feasible, the concordance of our findings with expected physiology demonstrates the potential utility of uEVs for future clinical applications. For example, uEV profiles may be able to identify patients with or at risk for salt-sensitive blood pressure or predict responsiveness to specific antihypertensive medications, such as mineralocorticoid receptor antagonists or diuretics.

There are several notable strengths to this analysis, including the novel study population, the achieved separation in dietary sodium intake, and the comprehensive evaluation of physiologic sodium regulation. However, the findings should be interpreted in the context of several key limitations, including relatively small sample size, unmeasured compensatory mechanisms such as vasopressin, volume status, and local paracrine factors, and inherent methodological limitations to measuring low abundance uEV proteins. uEV content reflects two processes – cellular protein abundance and sorting proteins into an exosome – and is therefore not a direct measurement of cellular protein expression, though a strong correlation between kidney tissue abundance and uEV concentration has been described in rodents.^12^ There is also some discussion of whether CD9 concentration is modified by dietary sodium intake,^34,49^ however, this has not been definitely established and the currently accepted standard is to normalize.^8,10^ Finally, the large number of comparisons increases the risk of type I error, though the physiologic concordance of the findings strengthens the inference.

## Perspectives

In normotensive adults with cardiometabolic risk factors, dietary sodium modulation induced physiologically concordant, nephron-segment specific changes in uEV protein concentration across all nephron segments. These findings support uEVs as a non-invasive tool to assess dynamic renal sodium handling and regulatory physiology in humans.

## Novelty and Relevance

### What is New?

- Dietary sodium intake induced nephron-segment specific changes in uEV protein abundance among patients with normal kidney function and risk factors for hypertension. The changes in uEV concentration in each diet mirrored expected physiology, which has not been previously shown in a normotensive population.
- Changes in uEV concentrations between high-sodium and low-sodium diets were not correlated with changes in systolic blood pressure as we observed very little sodium sensitivity in this normotensive population.

### What Is Relevant?

- The concordance of the changes in uEV protein concentrations under low-sodium and high-sodium diets with known, expected renal physiology suggests that uEVs can be used as a non-invasive tool to probe renal sodium handling in normotensive adults.

### Clinical Implications?

- Future investigations may use uEV profiles to identify patients with or at risk for salt-sensitive blood pressure or predict responsiveness to specific antihypertensive medications, such as mineralocorticoid receptor antagonists or diuretics.

## Data Availability

All data produced in the present study are available upon reasonable request to the authors

## Acknowledgments

None.

## Non-standard Abbreviations and Acronyms

11β-HSD2: 11β-hydroxysteroid dehydrogenase-2
AQP: Aquaporin
ENaC: Epithelial sodium channel
HS: High sodium
KCNJ: Potassium inwardly rectifying channel, subfamily J
LS: Low sodium
MR: Mineralocorticoid receptor
NCC: Sodium-chloride cotransporter
NKCC: Sodium-potassium-2-chloride cotransporter
OXSR: Oxidative stress-responsive kinase
RAAS: Renin-angiotensin-aldosterone system
SGK: Serum and glucocorticoid-regulated kinase
SGLT: Sodium-glucose cotransporter
SSBP: Salt sensitive blood pressure
TRPV: Transient receptor potential cation channel, subfamily V
uEV: Urinary extracellular vesicle

## Supplemental Results

**Table S1.**
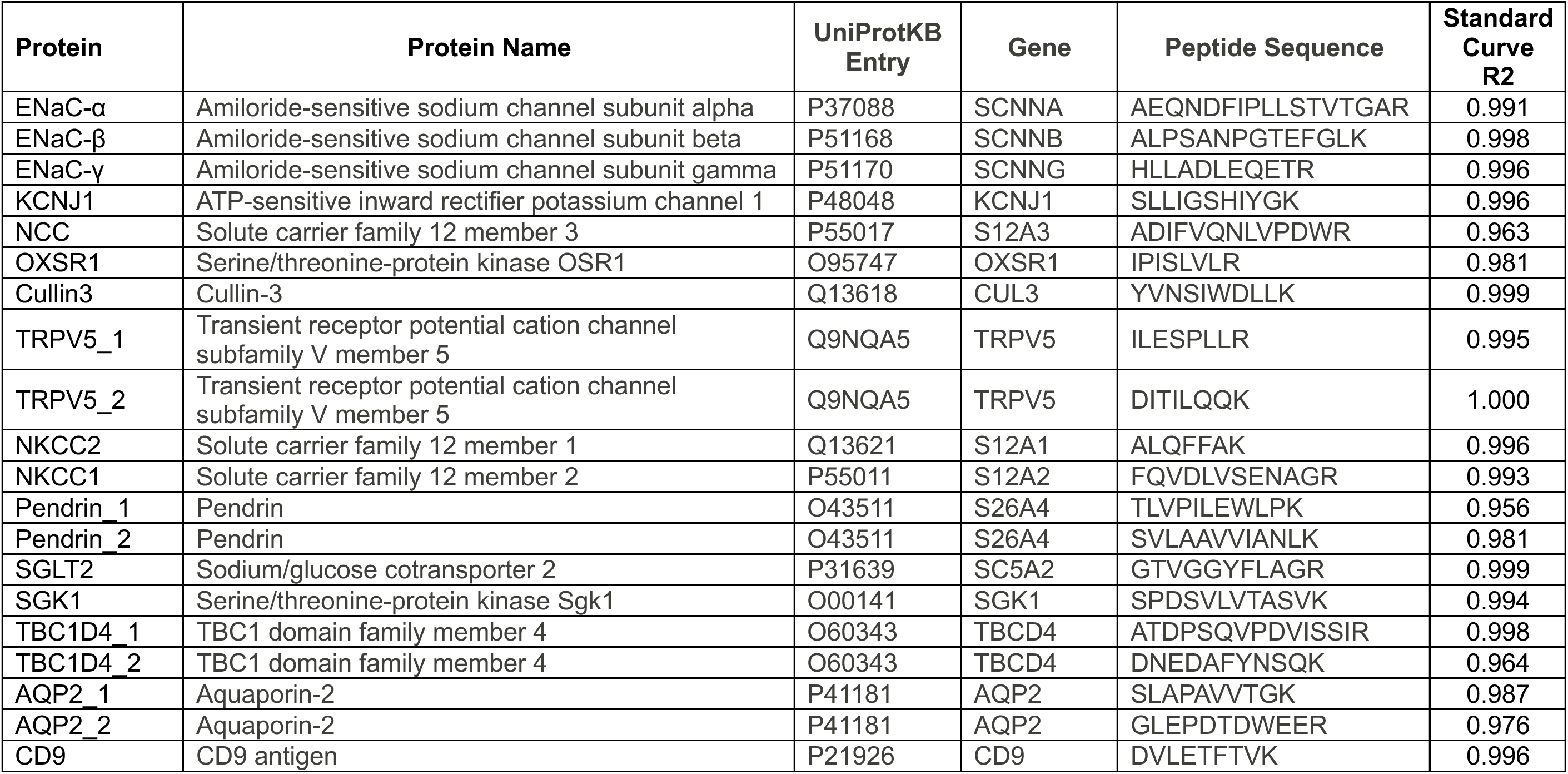
Peptide Standard and Results of Linearity Testing.

**Table S2.** Urine EV Concentration by Serum Aldosterone Tertile and Dietary Sodium Intake.

|  | **High Sodium Diet** |  |  |  |  | **Low Sodium Diet** |  |  |  |  |
| --- | --- | --- | --- | --- | --- | --- | --- | --- | --- | --- |
|  | **Tertile 1**<br>**4.5-8.4 ng/dl** | **Tertile 2**<br>**8.5-13.3 ng/dl** | **Tertile 3**<br>**13.4-42.9 ng/dl** | **KW *p*** | ***p*-**<br>**trend** | **Tertile 1**<br>**7.1-21.9 ng/dl** | **Tertile 2**<br>**21-28.9 ng/dl** | **Tertile 3**<br>**29-100 ng/dl** | **KW *p*** | ***p*-**<br>**trend** |
| NCC | 0.73 [0.61-1.74] | 0.87 [0.40-1.22] | 0.55 [0.41-0.91] | 0.46 | 0.50 | 1.04 [0.76-1.86] | 1.57 [1.06-1.94] | 0.80 [0.59-1.08] | 0.09 | 0.20 |
| ENaC-α | 0.01 [0.01-0.02] | 0.02 [0.01-0.03] | 0.02 [0.01-0.03] | 0.04 | 0.10 | 0.02 [0.01-0.02] | 0.02 [0.01-0.02] | 0.02 [0.01-0.03] | 0.42 | 0.49 |
| ENaC-β | 0.01 [0.01-0.02] | 0.01 [0.00-0.02] | 0.01 [0.01-0.02] | 0.81 | 0.73 | 0.01 [0.01-0.02] | 0.01 [0.01-0.02] | 0.01 [0.01-0.02] | 0.98 | 0.80 |
| ENaC-γ | 0.02 [0.01-0.03] | 0.02 [0.01-0.07] | 0.01 [0.00-0.02] | 0.22 | 0.38 | 0.03 [0.01-0.06] | 0.02 [0.02-0.03] | 0.02 [0.01-0.03] | 0.46 | 0.54 |
| ENaC combined | 0.03 [0.02-0.06] | 0.06 [0.03-0.13] | 0.05 [0.02-0.06] | 0.42 | 0.87 | 0.06 [0.03-0.11] | 0.06 [0.04-0.08] | 0.04 [0.03-0.08] | 0.50 | 0.86 |
| NKCC2 | 0.009 [0.003-0.01] | 0.01 [0.00-0.02] | 0.00 [0.00-0.01] | 0.17 | 0.80 | 0.00 [0.00-0.01] | 0.01 [0.00-0.01] | 0.01 [0.00-0.01] | 0.63 | 0.75 |
| NKCC1 | 1.04 [0.48-2.10] | 0.84 [0.59-1.16] | 0.65 [0.33-0.91] | 0.17 | 0.07 | 0.83 [0.52-1.63] | 0.99 [0.69-1.35] | 0.76 [0.62-1.29] | 0.76 | 0.33 |
| NKCC combined | 1.04 [0.49-2.11] | 0.87 [0.60-1.16] | 0.66 [0.37-0.92] | 0.19 | 0.09 | 0.84 [0.55-1.64] | 1.00 [0.70-1.36] | 0.77 [0.63-1.30] | 0.77 | 0.31 |
| TRPV5 1 | 0.08 [0.05-0.20] | 0.13 [0.07-0.29] | 0.10 [0.06-0.15] | 0.39 | 0.93 | 0.06 [0.02-0.15] | 0.04 [0.03-0.06] | 0.04 [0.03-0.06] | 0.60 | 0.57 |
| TRPV5 2 | 0.08 [0.05-0.17] | 0.11 [0.08-0.20] | 0.09 [0.04-0.15] | 0.46 | 0.99 | 0.07 [0.04-0.13] | 0.04 [0.04-0.05] | 0.05 [0.03-0.07] | 0.15 | 0.40 |
| TRPV5 combined | 0.17 [0.10-0.36] | 0.25 [0.15-0.47] | 0.21 [0.10-0.35] | 0.37 | 0.93 | 0.13 [0.07-0.30] | 0.08 [0.07-0.11] | 0.08 [0.06-0.13] | 0.30 | 0.42 |
| TBC1D4 1 | 0.01 [0.01-0.01] | 0.01 [0.01-0.02] | 0.01 [0.00-0.01] | 0.05 | 0.90 | 0.01 [0.00-0.01] | 0.01 [0.00-0.01] | 0.01 [0.005-0.02] | 0.96 | 0.62 |
| TBC1D4 2 | 0.006 [0.004-0.01] | 0.01 [0.00-0.04] | 0.00 [0.00-0.01] | 0.69 | 0.72 | 0.02 [0.00-0.03] | 0.01 [0.00-0.03] | 0.01 [0.003-0.01] | 0.12 | 0.29 |
| TBC1D4 combined | 0.01 [0.01-0.02] | 0.02 [0.01-0.08] | 0.02 [0.01-0.02] | 0.09 | 0.78 | 0.03 [0.01-0.05] | 0.03 [0.01-0.05] | 0.01 [0.01-0.03] | 0.31 | 0.47 |
| AQP2 1 | 2.13 [1.05-3.83] | 3.14 [1.47-5.27] | 1.73 [0.60-2.80] | 0.20 | 0.34 | 0.83 [0.55-1.11] | 1.17 [0.90-1.92] | 0.82 [0.59-1.69] | 0.25 | 0.89 |
| AQP2 2 | 3.29 [1.25-5.16] | 4.66 [1.96-6.03] | 2.33 [0.78-4.72] | 0.48 | 0.82 | 1.13 [0.77-2.41] | 1.41 [1.09-2.41] | 1.19 [0.78-3.21] | 0.80 | 0.52 |
| AQP2 combined | 5.87 [2.30-9.74] | 8.03 [5.02-13.31] | 4.08 [1.39-8.20] | 0.19 | 0.44 | 1.96 [1.34-3.80] | 3.33 [2.17-3.68] | 2.77 [1.51-4.52] | 0.74 | 0.93 |
| SGLT2 | 0.22 [0.15-0.28] | 0.22 [0.16-0.70] | 0.13 [0.09-0.21] | 0.06 | 0.18 | 0.22 [0.18-0.40] | 0.24 [0.18-0.37] | 0.20 [0.15-0.33] | 0.70 | 0.86 |
| OXSR1 | 0.06 [0.03-0.11] | 0.06 [0.05-0.12] | 0.05 [0.04-0.08] | 0.46 | 0.79 | 0.07 [0.03-0.22] | 0.07 [0.06-0.13] | 0.08 [0.06-0.13] | 0.71 | 0.55 |
| KCNJ1 | 0.08 [0.04-0.15] | 0.08 [0.04-0.18] | 0.06 [0.04-0.09] | 0.72 | 0.24 | 0.09 [0.08-0.41] | 0.09 [0.06-0.18] | 0.06 [0.05-0.10] | 0.12 | 0.14 |
| SGK1 | 0.01 [0.00-0.02] | 0.01 [0.01-0.05] | 0.01 [0.01-0.02] | 0.31 | 0.67 | 0.01 [0.01-0.02] | 0.02 [0.01-0.04] | 0.01 [0.01-0.02] | 0.03 | 0.12 |
| Pendrin | 0.21 [0.09-0.41] | 0.68 [0.10-1.19] | 0.19 [0.15-0.42] | 0.62 | 0.75 | 0.73 [0.54-1.38] | 0.76 [0.60-1.26] | 0.51 [0.38-1.07] | 0.34 | 0.30 |
| Cullin3 | 0.03 [0.01-0.07] | 0.05 [0.03-0.17] | 0.02 [0.01-0.03] | 0.03 | 0.54 | 0.03 [0.02-0.09] | 0.05 [0.02-0.07] | 0.02 [0.01-0.03] | 0.13 | 0.05 |
| Values represent median [IQR]; uEV concentrations normalized to CD9. Tertiles were calculated separately within each diet condition. KW, Kruskal-Wallis. *P*-trend is from a linear model, fit separately within HS and LS. |  |  |  |  |  |  |  |  |  |  |

**Table S3.**
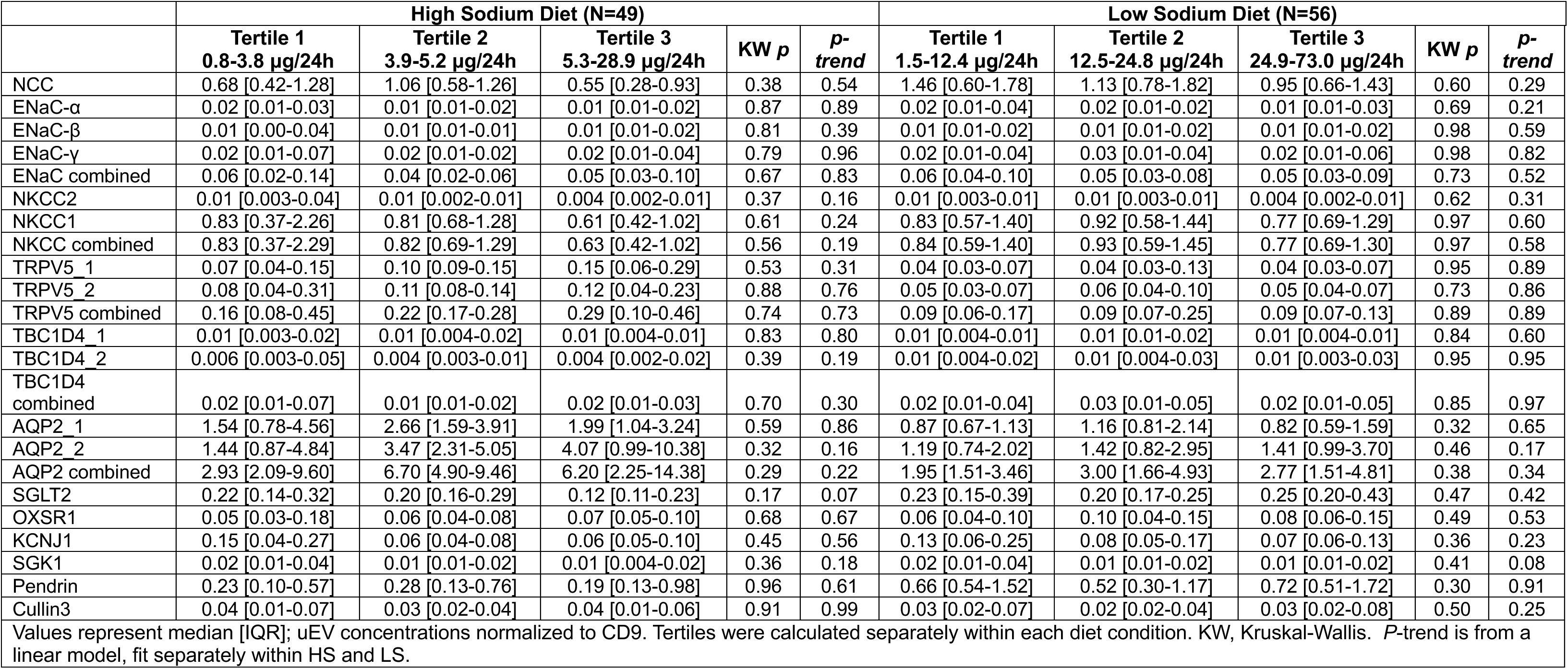
Urine EV Concentration by 24h Urine Aldosterone Tertile and Dietary Sodium Intake.

**Table S4.** Urine EV Concentration by Serum Cortisol-Cortisone Ratio Tertile and Dietary Sodium Intake.

|  | High Sodium Diet |  |  |  |  | Low Sodium Diet |  |  |  |  |
| --- | --- | --- | --- | --- | --- | --- | --- | --- | --- | --- |
|  | Tertile 1<br>1.1-4.2 | Tertile 2<br>4.3-5.5 | Tertile 3<br>5.6-11.3 | KW *p* | *p*-<br>trend | Tertile 1<br>2.9-4.3 | Tertile 2<br>4.4-5.1 | Tertile 3<br>5.2-12.8 | KW<br>*p* | *p*-<br>trend |
| NCC | 0.80 [0.36-1.23] | 0.68 [0.33-1.08] | 0.73 [0.55-1.40] | 0.77 | 0.31 | 0.89 [0.49-2.83] | 1.23 [0.82-1.80] | 1.05 [0.74-1.43] | 0.65 | 0.49 |
| ENaC-α | 0.02 [0.01-0.02] | 0.01 [0.01-0.03] | 0.01 [0.01-0.02] | 0.74 | 0.29 | 0.01 [0.01-0.02] | 0.02 [0.01-0.02] | 0.02 [0.01-0.03] | 0.81 | 0.64 |
| ENaC-β | 0.01 [0.01-0.01] | 0.01 [0.01-0.02] | 0.01 [0.01-0.02] | 0.57 | 0.31 | 0.01 [0.01-0.01] | 0.01 [0.01-0.02] | 0.01 [0.01-0.02] | 0.34 | 0.70 |
| ENaC-γ | 0.02 [0.01-0.03] | 0.02 [0.01-0.03] | 0.02 [0.01-0.03] | 0.82 | 0.39 | 0.02 [0.01-0.04] | 0.02 [0.01-0.04] | 0.02 [0.01-0.04] | 0.78 | 0.64 |
| ENaC combined | 0.05 [0.03-0.06] | 0.04 [0.02-0.08] | 0.05 [0.03-0.07] | 0.99 | 0.72 | 0.06 [0.03-0.09] | 0.05 [0.04-0.09] | 0.05 [0.03-0.08] | 0.78 | 0.37 |
| NKCC2 | 0.004 [0.002-0.01] | 0.01 [0.003-0.03] | 0.01 [0.002-0.01] | 0.12 | 0.38 | 0.01 [0.002-0.01] | 0.005 [0.003-0.01] | 0.005 [0.001-0.01] | 0.92 | 0.71 |
| NKCC1 | 0.54 [0.35-0.78] | 0.83 [0.49-1.32] | 1.02 [0.83-2.07] | 0.01 | 0.01 | 0.77 [0.55-1.32] | 0.86 [0.66-1.89] | 0.76 [0.68-1.16] | 0.69 | 0.18 |
| NKCC combined | 0.54 [0.36-0.79] | 0.84 [0.53-1.32] | 1.03 [0.84-2.08] | 0.01 | 0.01 | 0.77 [0.57-1.33] | 0.86 [0.66-1.90] | 0.76 [0.68-1.18] | 0.71 | 0.18 |
| TRPV5_1 | 0.11 [0.05-0.20] | 0.10 [0.06-0.17] | 0.09 [0.06-0.18] | 0.99 | 0.67 | 0.04 [0.03-0.07] | 0.05 [0.03-0.09] | 0.04 [0.03-0.06] | 0.46 | 0.57 |
| TRPV5_2 | 0.09 [0.03-0.16] | 0.09 [0.06-0.18] | 0.10 [0.06-0.21] | 0.67 | 0.19 | 0.05 [0.02-0.10] | 0.06 [0.04-0.07] | 0.04 [0.03-0.06] | 0.29 | 0.50 |
| TRPV5 combined | 0.20 [0.08-0.35] | 0.19 [0.12-0.42] | 0.20 [0.11-0.39] | 0.83 | 0.37 | 0.08 [0.06-0.21] | 0.10 [0.08-0.17] | 0.08 [0.06-0.11] | 0.35 | 0.51 |
| TBC1D4_1 | 0.01 [0.003-0.01] | 0.01 [0.01-0.02] | 0.01 [0.004-0.02] | 0.49 | 0.38 | 0.01 [0.01-0.01] | 0.01 [0.004-0.02] | 0.01 [0.005-0.02] | 0.96 | 0.79 |
| TBC1D4_2 | 0.004 [0.002-0.01] | 0.01 [0.003-0.03] | 0.01 [0.003-0.02] | 0.43 | 0.29 | 0.01 [0.003-0.03] | 0.01 [0.004-0.02] | 0.01 [0.005-0.03] | 0.82 | 0.65 |
| TBC1D4<br>combined | 0.01 [0.01-0.02] | 0.02 [0.01-0.05] | 0.01 [0.01-0.03] | 0.58 | 0.36 | 0.02 [0.01-0.04] | 0.02 [0.01-0.05] | 0.03 [0.01-0.05] | 0.72 | 0.59 |
| AQP2_1 | 2.05 [0.73-2.72] | 1.82 [1.19-4.28] | 3.10 [1.21-4.17] | 0.55 | 0.25 | 0.87 [0.61-1.23] | 1.22 [0.77-2.08] | 0.84 [0.57-1.09] | 0.25 | 0.62 |
| AQP2_2 | 2.74 [0.75-6.04] | 3.77 [1.52-5.33] | 2.69 [1.32-5.52] | 0.74 | 0.75 | 0.99 [0.52-1.76] | 2.30 [1.79-3.57] | 1.07 [0.82-1.37] | 0.01 | 0.49 |
| AQP2 combined | 5.44 [2.23-10.31] | 7.34 [3.20-10.62] | 4.75 [2.61-11.64] | 0.69 | 0.66 | 1.80 [1.04-3.56] | 3.58 [2.34-5.55] | 1.90 [1.57-2.87] | 0.02 | 0.92 |
| SGLT2 | 0.19 [0.13-0.26] | 0.20 [0.13-0.28] | 0.19 [0.12-0.53] | 0.90 | 0.22 | 0.21 [0.18-0.39] | 0.22 [0.15-0.38] | 0.24 [0.18-0.35] | 0.97 | 0.70 |
| OXSR1 | 0.06 [0.03-0.07] | 0.06 [0.04-0.11] | 0.06 [0.04-0.20] | 0.45 | 0.06 | 0.08 [0.05-0.13] | 0.08 [0.05-0.11] | 0.07 [0.04-0.17] | 0.88 | 0.83 |
| KCNJ1 | 0.06 [0.04-0.08] | 0.09 [0.05-0.17] | 0.06 [0.05-0.21] | 0.41 | 0.41 | 0.06 [0.05-0.20] | 0.07 [0.06-0.12] | 0.13 [0.06-0.19] | 0.62 | 0.78 |
| SGK1 | 0.01 [0.01-0.02] | 0.01 [0.01-0.03] | 0.01 [0.01-0.02] | 0.97 | 0.86 | 0.01 [0.01-0.02] | 0.01 [0.01-0.02] | 0.01 [0.01-0.03] | 0.84 | 0.68 |
| Pendrin | 0.20 [0.13-0.42] | 0.17 [0.09-0.59] | 0.41 [0.16-0.94] | 0.52 | 0.66 | 0.72 [0.49-1.67] | 0.80 [0.56-1.58] | 0.51 [0.27-0.64] | 0.05 | 0.06 |
| Cullin3 | 0.03 [0.01-0.06] | 0.03 [0.02-0.05] | 0.04 [0.02-0.08] | 0.77 | 0.71 | 0.03 [0.02-0.07] | 0.02 [0.02-0.05] | 0.03 [0.02-0.05] | 0.89 | 0.20 |

Values represent median [IQR]; uEV concentrations normalized to CD9. Tertiles were calculated separately within each diet condition. KW, Kruskal-Wallis. *P*-trend is from a linear model, fit separately within HS and LS.

**Figure S1.**
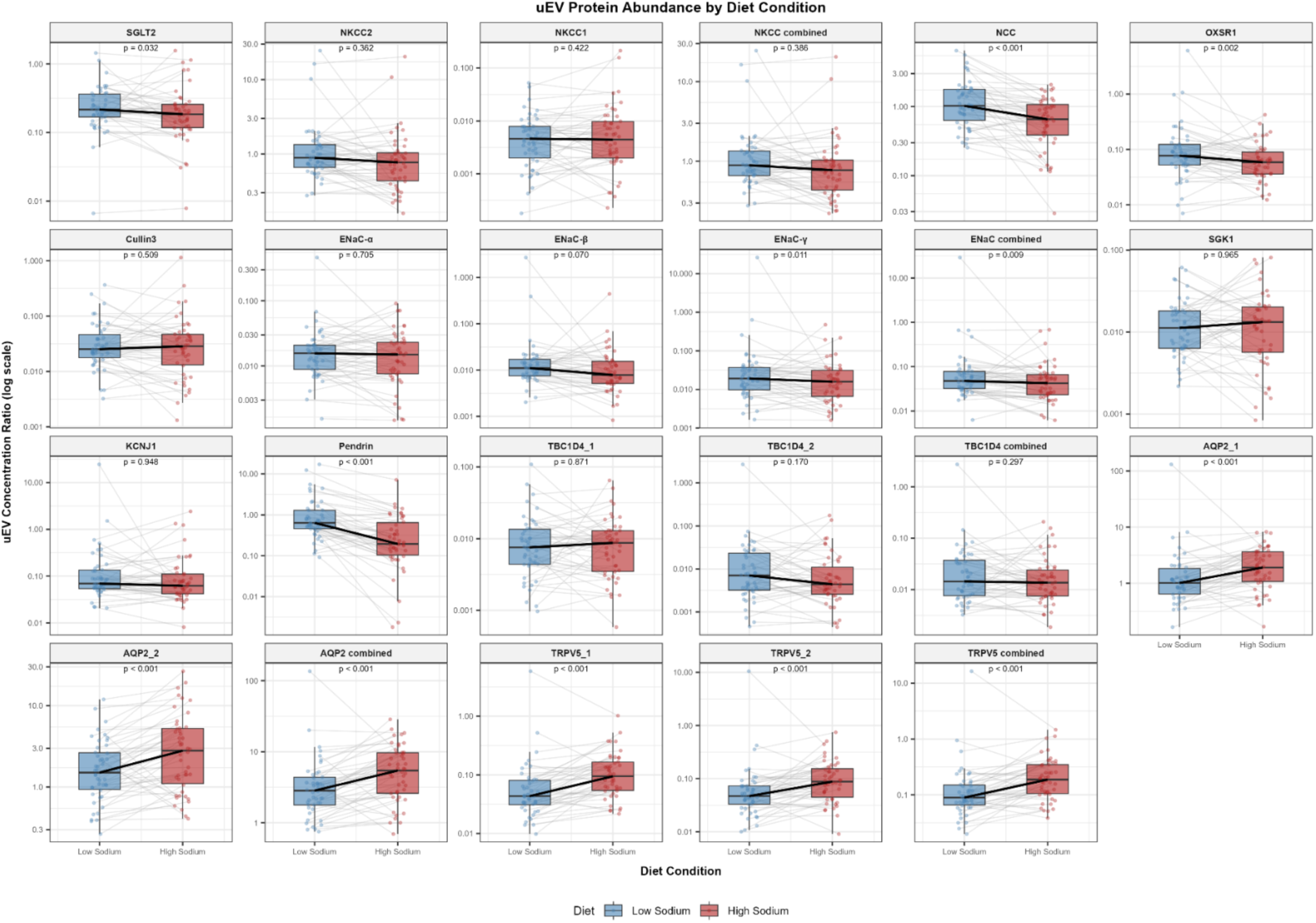
Boxplots summarizing the distribution of uEV concentration by diet, among matched pairs (N=46). Boxplots show the median (center line), interquartile range (box), and whiskers extending to the most extreme values within 1.5x the interquartile range. Individual observations are overlaid. Data are displayed on a log_10_ scale. Thin gray lines connect paired values from the same participant. Solid black line connects the median across all participants. P-values from Wilcoxon signed rank.

**Figure S2.**
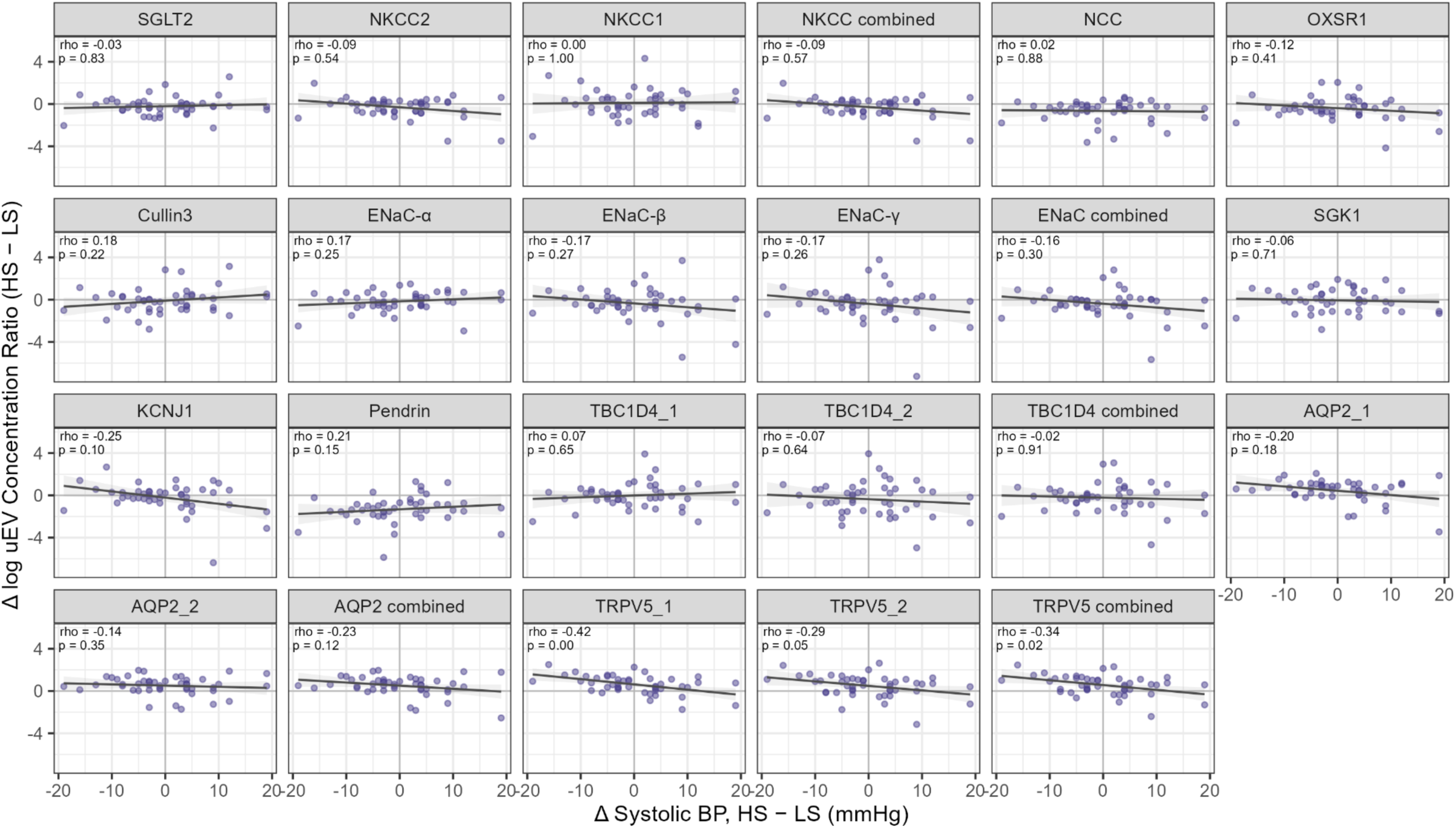
Scatterplots summarizing the correlation between change in uEV concentration and change in systolic blood pressure across diet conditions, among matched pairs (N=46). Both differences are calculated as high sodium minus low sodium, such that positive values indicate a higher value during high sodium intake. Change in uEV concentration is expressed as the difference in log-transformed values. Scatterplots show individual observations. The trend line is the ordinary least-squares regression fit and the shaded band is the 95% confidence interval around the fitted line. Spearman’s rho and the associated p-value are overlaid in each panel.

**Figure S3.**
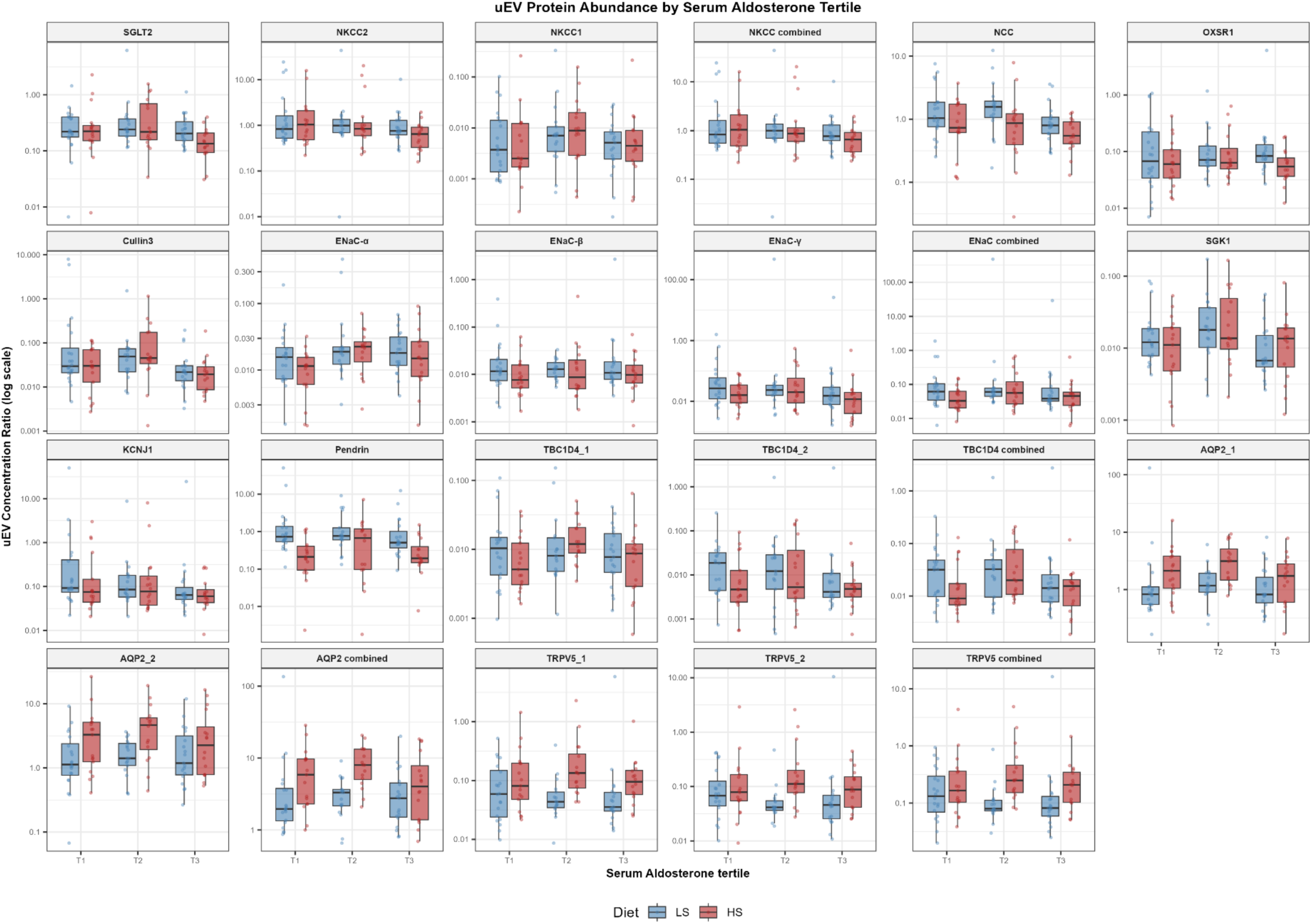
Boxplots summarizing the distribution of uEV concentration by diet-specific serum aldosterone tertile. Boxplots show the median (center line), interquartile range (box), and whiskers extending to the most extreme values within 1.5x the interquartile range. Individual observations are overlaid; all participants included (Low Sodium (LS) N=56, High Sodium (HS) N=49). Data are displayed on a log_10_ scale. Tertiles calculated separately for HS and LS diet within the full cohort.

**Figure S4.**
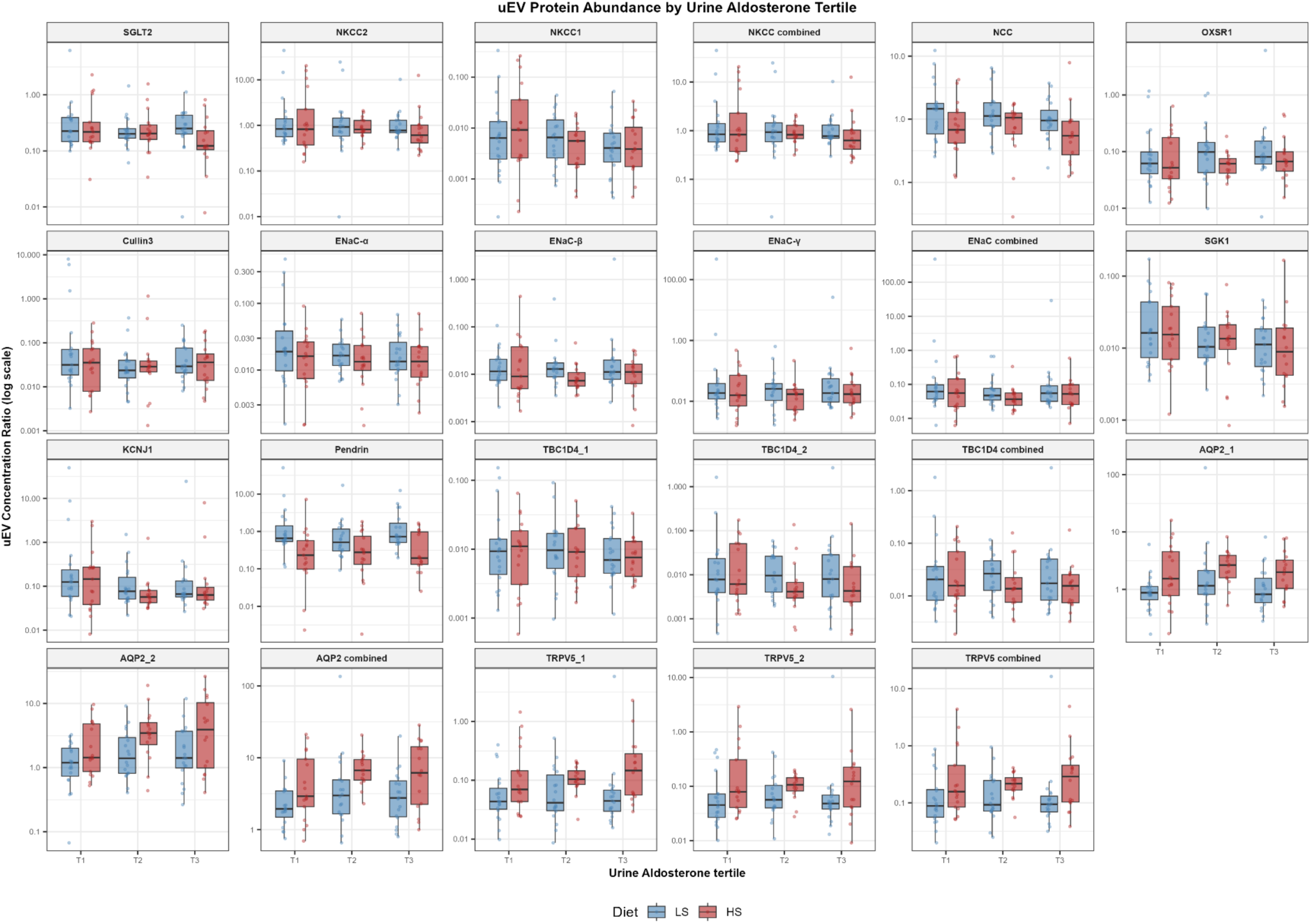
Boxplots summarizing the distribution of uEV concentration by diet-specific urine aldosterone tertile. Boxplots show the median (center line), interquartile range (box), and whiskers extending to the most extreme values within 1.5x the interquartile range. Individual observations are overlaid; all participants included (Low Sodium (LS) N=56, High Sodium (HS) N=49). Data are displayed on a log_10_ scale. Tertiles calculated separately for HS and LS diet within the full cohort.

**Figure S5.**
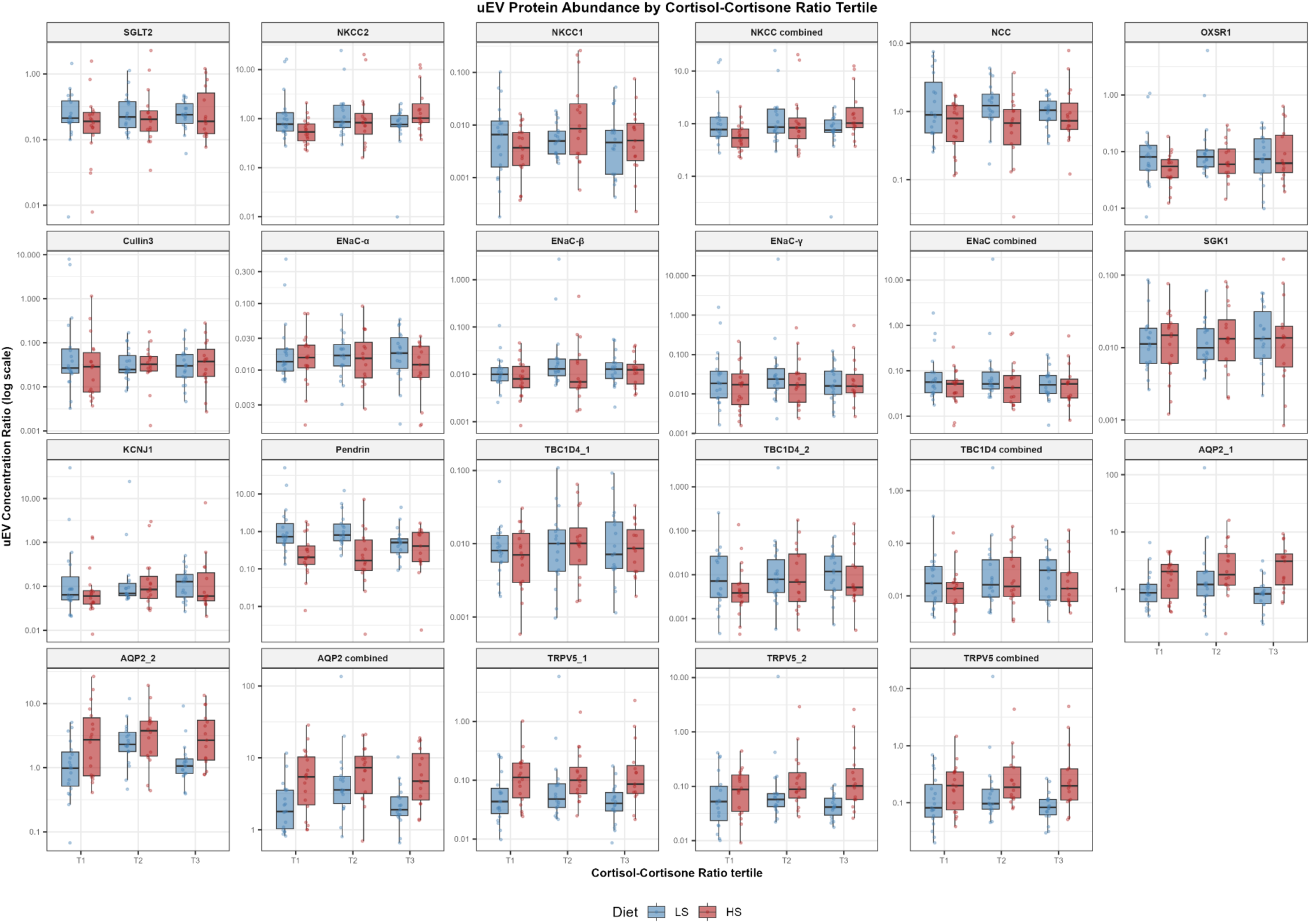
Boxplots summarizing the distribution of uEV concentration by diet-specific cortisol-cortisone ratio. Boxplots show the median (center line), interquartile range (box), and whiskers extending to the most extreme values within 1.5x the interquartile range. Individual observations are overlaid; all participants included (Low Sodium (LS) N=56, High Sodium (HS) N=49). Data are displayed on a log_10_ scale. Tertiles calculated separately for HS and LS diet within the full cohort.

